# Leveraging Artificial Intelligence to Mitigate Adolescent Risky Behaviors: A Scoping Review Protocol

**DOI:** 10.1101/2024.06.11.24308749

**Authors:** Hamidreza Sadeghsalehi, Hassan Joulaei

## Abstract

Adolescents are particularly vulnerable to engaging in risky behaviors such as violence, unprotected sex, and substance abuse, which have significant negative impacts on their health and development. Recent advancements in artificial intelligence (AI) offer innovative solutions to address these behaviors, yet the evidence regarding the efficacy and implementation of AI-based interventions remains fragmented. This scoping review aims to systematically explore and map the literature on AI-based interventions designed to reduce risky behaviors among adolescents.

This review will follow the methodological frameworks outlined by Arksey and O’Malley (2005) and improved by Levac, Colquhoun, and O’Brien (2010), in line with the Joanna Briggs Institute guidelines. The PRISMA Extension for Scoping Reviews (PRISMA-ScR) will guide the reporting. The search strategy will be executed across PubMed, Scopus, Web of Science Core Collection, CINAHL, PsycINFO, Cochrane Central Register of Controlled Trials, Embase, SID, and Magiran, focusing on articles published up to June 2024 in English and Farsi. Titles and abstracts will be screened by two independent reviewers using Rayyan, followed by full-text screening of relevant studies. Data will be charted using a standardized form, and discrepancies will be resolved through discussion or by consulting a third reviewer. Data will be synthesized descriptively and presented in tables, figures, and diagrams.

## Introduction

### Background and Rationale

Adolescence is a transformative period marked by significant physical, emotional, and social changes (1). This stage of life is characterized by a heightened propensity for engaging in risky behaviors, such as violence, unprotected sex, and substance abuse. These behaviors pose significant threats to adolescents’ health and well-being, often leading to immediate and long-term adverse outcomes such as injuries, chronic health conditions, and mental health issues (2-6). Despite various traditional interventions aimed at curbing these behaviors, the prevalence of risky behaviors among adolescents remains a major public health concern globally (7).

Artificial Intelligence (AI), a rapidly evolving field encompassing machine learning, predictive modeling, and data analysis, offers promising tools for addressing complex health issues (8, 9). AI is capable of analyzing huge datasets to find trends and related risk factors associated with adolescent risky behaviors, enabling early detection and tailored interventions (10, 11). The integration of AI in health interventions holds potential for improving the effectiveness of strategies designed to reduce risky behaviors in adolescents.

Adolescents today are increasingly engaged with the internet and social media, which have become integral parts of their daily lives. Teenagers have access to smartphones in 95% of cases, and 45% of them are online nearly all the time, according to the Pew Research Center (12). Social media platforms, such as Instagram, Snapchat, and TikTok, are particularly popular, with a majority of teenagers using these platforms regularly (13). This high level of internet and social media engagement exposes adolescents to various risks, including cyberbullying, exposure to inappropriate content, and peer pressure, which can contribute to risky behaviors (14). Given this context, leveraging AI to monitor and mitigate these risks becomes increasingly important. AI technologies can analyze social media interactions and online activities to identify early warning signs of risky behaviors and provide timely interventions (15). By harnessing AI’s capabilities in real-time data analysis and pattern recognition, there is significant potential to enhance the effectiveness of preventive measures and support adolescents in navigating the challenges of the digital age more safely.

However, the application of AI in this domain is still emerging, and there is a need to map the existing evidence to understand its current state, identify gaps, and outline future research directions (16). While there have been reviews on AI applications in healthcare broadly, a focused review on its use specifically for reducing adolescent risky behaviors is lacking. This gap highlights the need for a comprehensive scoping review to systematically explore and synthesize the available evidence.

A scoping review is justified over other types of reviews, such as systematic reviews or meta-analyses, due to the nascent and exploratory nature of the research topic (17). Scoping reviews are particularly useful for examining emerging areas, identifying key concepts, and mapping the extent of research activity. They provide an overview of the available evidence without necessarily assessing the quality of included studies or synthesizing findings in a manner typical of systematic reviews (18).

Existing literature on AI applications in adolescent health primarily focuses on broader health outcomes rather than specifically targeting risky behaviors. For instance, AI has been explored in mental health interventions (19-21), chronic disease management (22, 23), and general preventive health strategies (24, 25). However, a dedicated review on how AI is being utilized to mitigate risky behaviors in adolescents is absent. This scoping review aims to fill this gap by providing a detailed overview of the current landscape of AI interventions targeting adolescent risky behaviors, thereby offering insights into the opportunities and challenges in this field.

### Objective

The main goal of this scoping review is to methodically investigate and map the current body of literature on AI-based interventions aimed at reducing risky behaviors among adolescents. Specifically, the review will address the following research questions:

What types of AI-based interventions have been implemented to reduce risky behaviors in adolescents?

What are the outcomes of these AI-based interventions in terms of reducing risky behaviors?

What are the key challenges and opportunities associated with the implementation of AI-based interventions in this context?

What gaps exist in the current research on AI interventions targeting adolescent risky behaviors?

By tackling these inquiries, the aim of this scoping review is to present a comprehensive overview of the application of AI in reducing adolescent risky behaviors and identify directions for future research and intervention development.

## Methods

This scoping review will follow the methodology outlined by Arksey and O’Malley (2005)(26), with enhancements by Levac, Colquhoun, and O’Brien (2010)(27). The review protocol has been developed following the Joanna Briggs Institute (JBI) guidelines for conducting scoping reviews (28). The PRISMA Extension for Scoping Reviews (PRISMA-ScR) will be used as the reporting guideline for the scoping review manuscript to ensure comprehensive and transparent reporting of the findings (29).

### Eligibility Criteria

#### Inclusion Criteria

**Population**: Adolescents aged 10-19 years.

**Intervention**: AI-based interventions (e.g., machine learning, deep learning) aimed at reducing risky behaviors.

**Comparison**: Traditional interventions or no intervention.

**Outcomes**: Reduction in risky behaviors (e.g., substance abuse, violence, unprotected sex, smoking).

**Study Types**: All original study designs, including RCTs, cohort studies, cross-sectional studies, case-control studies, and qualitative studies.

**Language**: Articles published in English and Farsi.

**Publication Date**: Studies published up to June 2024.

#### Exclusion Criteria

**Population**: Studies focusing on children below 10 years or adults above 19 years.

**Intervention**: Non-AI-based interventions.

**Outcomes**: Studies that do not measure risky behaviors.

**Study Types:** Review studies.

**Language**: Articles published in languages other than English and Farsi.

#### The rationale for Criteria

Adolescents are targeted due to the unique developmental challenges and propensity for risky behaviors in this age group.

AI-based interventions are the focus due to their potential for innovative solutions.

The inclusion of various study designs ensures a comprehensive understanding of the evidence landscape.

Limiting the review to articles in English and Farsi and from the past 13 years ensures the inclusion of relevant and accessible studies.

#### Limitations/Restrictions

Exclusion of non-English and non-Farsi studies may result in language bias.

#### Handling Ambiguous Information

Ambiguous information will be discussed among the review team to reach a consensus. In cases where consensus is unattainable, a third reviewer will be brought in for further evaluation.

#### Shared Interpretation of Criteria

Regular team meetings will be held to ensure all members have a consistent understanding of the eligibility criteria.

### Information Sources and Search Strategy

#### Sources to be Searched

**Databases:** PubMed, Scopus, Web of Science Core Collection, CINAHL, APA PsycINFO, Cochrane Central Register of Controlled Trials (CENTRAL), Embase, SID, and Magiran

**General Search Terms:** “artificial intelligence,” “machine learning,” “deep learning,” “adolescents,” “teenagers,” “risky behaviors,” “substance abuse,” “violence,” “unprotected sex,” “alcohol drinking,” “smoking”

#### Citation Management Software

EndNote will be used for managing citations.

#### PRESS Checklist

The PRESS (Peer Review of Electronic Search Strategies) checklist will be used to ensure the quality and comprehensiveness of the search strategy.

### Study Selection/Screening

#### Eligibility Criteria for Screening

The same inclusion and exclusion criteria will be employed for the screening process.

#### Title and Abstract Screening

The title and abstract of all identified studies will be screened independently by two reviewers. A pilot screening of a sample of articles will be conducted to ensure consistency.

Discrepancies will be resolved through discussion, and if necessary, a third reviewer will be consulted.

Rayyan will be used to manage and screen articles.

#### Full-Text Screening

Full-text articles of potentially relevant studies will be retrieved and assessed by two reviewers independently.

Discrepancies will be addressed through dialogue or by seeking input from a third reviewer.

Rayyan will be used to manage and screen full-text articles.

### Data Charting/Collection/Extraction

#### Data Charting Form Development

A standardized data charting form will be developed based on the JBI guidelines.

#### Data Items to be Collected

Study characteristics (e.g., author, year, country) Population (e.g., age, gender)

Intervention details (e.g., type of AI, specific intervention) Outcomes (e.g., type of risky behavior, outcome measures) Key findings

Challenges and limitations

#### Storage of Rules for Data Extraction

Detailed rules for data extraction will be documented and shared among the review team to ensure consistency.

#### Data Extraction Process

Data will be extracted independently by two reviewers. A pilot extraction will be conducted on a sample of articles. In order to resolve any discrepancies, discussion or consultation with a third reviewer will be undertaken.

Unclear or missing information will be addressed by contacting the study authors if necessary.

#### Handling Friend Studies

Studies conducted by the same research team or with overlapping data will be carefully examined to avoid double-counting results.

### Synthesis and Presentation of Results

#### Data Cleaning

Data will be cleaned and organized using Microsoft Excel.

#### Software for Data Cleaning and Analysis

Microsoft Excel will be used for data cleaning and qualitative analysis, respectively.

#### PRISMA Flow Diagram

A PRISMA flow diagram will be used to illustrate the study selection process.

#### Data Synthesis

Data will be synthesized descriptively. The analysis will include the frequency and percentage of studies in each category, trends in publication over time, and comparison of the number of studies across different countries.

Findings will be presented in tables, figures, and diagrams to provide a comprehensive overview.

#### Presentation of Data Items

Each data item will be presented in a meaningful way, with the support of stakeholders where necessary, to ensure the results are relevant and actionable.

### Ethics and Dissemination

#### Ethics Approval

Ethics approval is not required for this scoping review as it involves secondary data analysis of published studies.

#### Roles of Collaborators/Stakeholders

Collaborators and stakeholders will be involved in the development of the search strategy, data extraction, and interpretation of findings.

#### Dissemination Plans

The outcomes of this scoping review will be distributed through peer-reviewed publications, conference presentations, and stakeholder meetings to ensure wide reach and impact.

This comprehensive methodology ensures a rigorous and systematic approach to mapping the landscape of AI interventions aimed at reducing risky behaviors in adolescents.

## Conclusion

The main goal of this scoping review is to methodically investigate and map the current body of literature on AI-based interventions aimed at reducing risky behaviors among adolescents. The secondary objectives are to identify the types of AI-based interventions implemented, evaluate their outcomes in terms of reducing risky behaviors, examine the challenges and opportunities associated with their implementation, and identify gaps in the current research on this topic.

This scoping review intends to fill several critical gaps in the existing literature. First, it will provide a comprehensive overview of the current landscape of AI interventions targeting adolescent risky behaviors, an area that has not been extensively reviewed. Second, by identifying the outcomes and effectiveness of these interventions, the review will offer valuable insights into which strategies are most promising. Third, it will highlight the key challenges and opportunities in implementing AI-based interventions, providing guidance for future research and practical application. Lastly, by mapping the gaps in the current research, the review will outline areas that require further investigation, thereby helping to shape future studies and interventions. Advancing the field, this scoping review will serve as a foundational resource for researchers, practitioners, and policymakers interested in leveraging AI to address risky behaviors in adolescents. By synthesizing the existing evidence, it will inform the development of more effective, targeted, and scalable interventions. Additionally, the review’s findings will help to guide funding priorities and policy decisions aimed at enhancing adolescent health outcomes through innovative AI solutions.

## Data Availability

All data produced in the present work are contained in the manuscript.

